# Variation in hospital cost trajectories at the end of life by age, multimorbidity and cancer type

**DOI:** 10.1101/2022.02.22.22271323

**Authors:** Katharina Diernberger, Xhyljeta Luta, Joanna Bowden, Joanne Droney, Elizabeth Lemmon, Giovanni Tramonti, Bethany Shinkins, Ewan Gray, Joachim Marti, Peter S Hall

## Abstract

**Background:** Approximately thirty thousand people in Scotland are diagnosed with cancer annually, of whom a third live less than one year. The timing, nature and value of hospital-based healthcare for patients with advanced cancer are not well understood. The aim of this study was to describe patterns of hospital-based healthcare use and associated costs in the last year of life for patients with a cancer diagnosis.

**Methods:** We undertook a Scottish population-wide administrative data linkage study of hospital-based healthcare use for individuals with a cancer diagnosis aged 60 years and over on their date of death, who died between 2012 and 2017. Hospital admissions, length of stay (LOS), number and nature of outpatient and day case appointments were analysed for all cancer types. Generalised linear models were used to adjust costs for age, gender, socioeconomic deprivation status, rural-urban (RU) status and comorbidity.

**Results:** The study included 85,732 decedents with a cancer diagnosis, for whom 64,553 (75.3%) cancer was the primary cause of death. Mean age at death was 80.01 (SD 8.15) years. The mean number of inpatient stays in the last year of life was 5.88 (SD 5.68), with a mean LOS of 7 days. Mean total inpatient, outpatient and daycase costs per patient were £10261, £1275 and £977 respectively. Admission rates rose sharply in the last month of life. One year adjusted and unadjusted costs decreased with increasing age. A higher comorbidity burden was associated with higher costs and major cost differences between cancer types were also observed.

**Conclusions:** People in Scotland in their last year of life with cancer are high users of secondary care. Hospitalisation accounts for a high proportion of costs, particularly in the last month of life. Further research is needed to examine triggers for unplanned hospitalisation and to identify modifiable reasons for variation in hospital use among different cancer cohorts.

## Background

Approximately thirty thousand people in Scotland are diagnosed with cancer each year, of whom 10,000 live less than one year. (ISD-Scotland, 2022) It is estimated that two out of five people will develop cancer in their lifetime. Over the last decade, cancer incidence has risen in Scotland, whilst the mortality rate has fallen. This trend can be explained by improvements in diagnosis and the development of newer anti-cancer therapies, the ageing population and the fact that cancer incidence increases with age. (Scottish Government, 2021)

In 2018 there were 16,153 cancer deaths registered in Scotland, excluding non-melanoma skin cancers. (ISD-Scotland, 2019) A quarter of all deaths from cancer (n=3,980) were attributed to lung cancer, followed by colorectal (n=1,743), breast (n=1,001), prostate (n=923), and oesophageal (n=873) cancers. These five cancer types were responsible for more than half of the Scottish cancer deaths.

People who are nearing the end of life are high users of secondary care services. (Diernberger, et al., 2021) Currently around 50% of people in Scotland die in hospital. (Bekelman, et al., 2016; Clark, et al., 2014; NRS, 2020) A recent paper described trends in place of death in Scotland between 2004 and 2016, finding a reduction in hospital deaths, from 58% to 50.1%, during the study period along with a corresponding increase in deaths in community settings including care homes. (Finucane, et al., 2019)

Hospitalisation of patients in the last year of life may be recommended and necessary for some people with complex clinical needs and increasing proximity to death. Nevertheless, evidence suggests that clinical interventions close to the end of life may also represent a clinical culture of ‘over-medicalisation’, with limited or no meaningful benefit to individuals. (Mills, Buchanan, Guthrie, Donnan, & Smith, 2019; Hughes-Hallet, Craft, Davies, Mackay, & Nielsson, 2011; Earle, et al., 2004) It is increasingly accepted that we should align clinical intervention with individual patients’ needs and preferences, moving away from the historic ‘doctor knows best’ culture. (Realistic Medicine, 2022)

The rising costs of cancer treatment, driven by new therapeutic options, is important context and necessitates that the true value of clinical interventions is understood so that scarce resources can be directed appropriately. (Vokinger, 2020; Diernberger, K. et al. 2021)

A systematic review, which included all English language retrospective studies looking at cost in cancer care using administrative data, showed that a range of sociodemographic, clinical and health system characteristics influenced costs. Further outcomes reported an exponential cost increase with proximity to death and showed inpatient care as the main driver of this. (Langton, et al., 2014) A systematic review of Scotland-based palliative care research published in 2018 revealed a lack of health economic research. (Finucane, et al., 2018)

Our recent study of secondary care costs for end of life care included the Scottish population who died between 2012 and 2017. We showed that intensity of healthcare use and costs were highest in cancer patients, mainly due to inpatient stays. (Diernberger, et al., 2021) Similar results were found in our English parallel study. (Luta, et al., Healthcare trajectories and costs in the last year of life: a retrospective primary care and hospital analysis., 2020) Furthermore, in both studies the cancer cohort demonstrated a particularly steep cost-increase in the final three months of life, again largely as a result of inpatient hospitalisation.

In the present study, we sought to understand more about the timing and nature of secondary healthcare use for patients with (advanced) cancer in their last year of life in Scotland, to identify factors associated with variation and to question the extent to which current care pathways offer value.

## Methods

The study population in this retrospective cohort analysis included everyone with a recorded cancer diagnosis in Scotland who died between 2012 and 2017 and were over 60 years of age on their date of death. Via the linkage of cancer records (SMR06) to data from Scottish hospital records, hospital-based healthcare use over the last twelve months of life was examined. The final dataset included cancer registry data, inpatient, outpatient and day-case activity (SMR00 and SMR01) and the National Records of Scotland (NRS) death registration data.

### Data sources

Data were obtained via the Scottish Research Data Safe Haven from Public Health Scotland, who manage all health related data connected to NHS Scotland. Linkage was established using the Community Health Index (CHI) number as the primary key. (NHS, 2021) The Scottish Morbidity Record (SMR) outpatient (SMR00), inpatient and day case (SMR01) and the National Records of Scotland (NRS) record of deaths were linked to cancer registry data (SMR06). SMR01 includes episode-based patient records that relate to all acute inpatient and day cases. SMR00 relates to all outpatient activity including new and follow-up appointments. NRS manages the official register for deaths in Scotland, which includes all deaths with details on causes on death from a death certificate. All patient identifiers including the CHI were removed from the datasets prior to release in the National Safe Haven.

### Inclusion and exclusion criteria

Data linkage and detailed eligibility criteria are reported in Figure S1 (supplementary material). Major inclusion criteria were:

- Death registered between January 1^st^ 2012 and December 31^st^ 2017
- Age at death ≥60 years
- Healthcare data available for a minimum of 365 days prior to death
- A linked record available in the cancer registry between 1^st^ 2012 and December 31^st^ 2017

In the selection process of the study population, the NRS death dataset of the eligible cohort of decedents was merged with the outpatient (SMR00) and the inpatient and day-case (SMR01) dataset. Inpatient and outpatient resource-use data was excluded if the patient identifier (PID) was missing or if the resource use occurred outside the study period. Following this, SMR06 data was merged onto the existing clean dataset and decedents who did not have a cancer diagnosis were excluded. Data of decedents with a cancer diagnosis, fulfilling all other criteria for inclusion, were retained for the final analysis.

### Patient characteristics

Patient characteristics included gender, age and primary cause of death, with a subsequent division into cancer as the primary cause of death and an “others” category. Cancer types were grouped based on number of patients and/or using the first two digits of the ICD-10 code. Comorbidity was estimated using the Charlson Comorbidity Index (CCI),based on secondary care coding, which entailed a 5 year look back from patients’ first contact with secondary care. (Pylväläinen, et al., 2019) A rural-urban indicator was included, as was the Scottish index of Multiple Deprivation (SIMD). (Scottish-Government, 2020; SIMD, 2022)

### Outcome measures

#### Inpatient and Day Care

Hospital inpatient care in the last year of life was captured as the number of hospital admissions, the timing of these in relation to death, the mean number of bed days per inpatient stay and the total number of bed days over the twelve-month period.

Scottish health service costs (Scottish cost book) were used to estimate the cost of inpatient care, mainly R040 (Specialty group costs including inpatient data for all specialties excluding long stays) and R040LS to include more detail regarding long inpatient stays. (Public Health Scotland, 2019) Critical care stays were included within mean costs. Day cases were costed using R042 (Specialty group costs for day cases).

#### Outpatient care

Hospital outpatient data included the number and nature of outpatient appointments per patient in the last year of life. Costs for outpatient appointments were derived from the Scottish health service costs documents R044 (Specialty group costs for consultant led outpatient appointments), R045 (Specialty group costs for nurse led clinics) and R046 (Specialty group costs for Allied Health Professionals services). The costs were based on national average unit costs for each service code.

### Statistical Analysis

Descriptive statistics were used to characterise the study population. Means and standard deviation (SD) were calculated for service use and costs. Generalised linear models (GLM) as recommended by Glick et al. (2014) were used to model costs as they are robust to the skewed distributions typical for healthcare related cost data. Known important predictors of costs are age, gender, primary cause of death, deprivation, urban-rural indicator and comorbidity. (Hazra, Rudisill, & Gulliford, 2018; Moran, Solomon, Peisach, & Martin, 2007) The effect of age, primary cause of death and CCI were estimated in isolation, with the other predictors included as covariates in the GLM. Potential interactions between age and gender as well as age and cause of death were also assessed. Analysis was carried out using Stata version 16 (StataCorp, College Station, TX, USA).

## Results

### Patient characteristics

Table 1 displays the patient characteristics for the final cohort comprising 85,732 decedents with a cancer diagnosis. Slightly over half of the study population was male (52.24%), and the greatest proportion of decedents were aged between 70 and 84 years at time of death. The most common cancer type as a primary cause of death (by three-fold) was lung cancer, making up over 20% of the included population.

**Table 1:** Patient characteristics

|  | Frequency | Percent |
| --- | --- | --- |
| Male | 44787 | 52.24 |
| Female | 40945 | 47.76 |
| 60-64 | 11937 | 13.92 |
| 65-69 | 14640 | 17.08 |
| 70-74 | 17279 | 20.15 |
| 75-79 | 17594 | 20.52 |
| 80-84 | 14173 | 16.53 |
| 85-89 | 7817 | 9.12 |
| 90+ | 2292 | 2.67 |
| Cancer not main cause of death | 21179 | 24.70 |
| Bronchus/Lung | 18372 | 21.43 |
| Colon/Rectosigmoideum/Rectum | 6342 | 7.40 |
| Oesophagus/Stomach | 5540 | 6.46 |
| Kidney/Bladder | 3555 | 4.15 |
| Liver/Intrahepatic | 2398 | 2.80 |
| Pancreas | 3334 | 3.89 |
| Haematological | 2342 | 2.73 |
| Brain | 1158 | 1.35 |
| Breast | 2136 | 2.49 |
| Ovary | 1331 | 1.55 |
| Prostate | 2846 | 3.32 |
| "other" cancer | 15199 | 17.73 |
| Comorbidity Index (CI) 0 or not recorded | 20325 | 23.71 |
| CI 1- CI 3 | 53412 | 62.30 |
| CI 4 - CI 6 | 5143 | 6.00 |
| CI 7 - CI 12 | 6852 | 7.99 |
| SIMD 1st quintile (Most deprived) | 19393 | 22.64 |
| SIMD 2<sup>nd</sup> quintile | 19649 | 22.94 |
| SIMD 3<sup>rd</sup> quintile | 17876 | 20.87 |
| SIMD 4<sup>th</sup> quintile | 15260 | 17.82 |
| SIMD 5th quintile (Least deprived) | 13462 | 15.72 |
| 1 - Large urban area | 37967 | 44 |
| 2 - Other Urban Areas | 30263 | 35.30 |
| 3 - Accessible Small Towns | 8194 | 9.56 |
| 4 - Remote Small Towns | 2258 | 2.63 |
| 5 - Accessible Rural Areas | 1288 | 1.50 |
| 6 - Remote Rural Areas | 5762 | 6.72 |

Cancer was recorded as the primary cause of death in 64,553 (75.3%) patients. Patients who had a cancer diagnosis and died from cancer tended to be younger compared to those with another cause of death. For more details see supplementary Figure 2.

Table 2 presents patient characteristics split by cancer type. The first column presents the cohort of patients who had a cancer diagnosis regardless of the main cause of death. The second column consists solely of those with cancer as the main cause of death. Compared to the whole cohort, patients in this category were slightly younger, from more deprived areas and had a higher level of comorbidity. Despite the study population being limited to older adults (60+), we observed differences in age at death across cancer types, with prostate cancer patients being oldest at their time of death. The highest comorbidity burden was detected in those dying from ovarian cancer. There were noticeable differences in the socioeconomic (SIMD) and rural-urban status of patients by cancer type. Patients who died from lung cancer as their main cause of death were found to have the lowest SIMD with a mean value of 2.53 (1.33) whilst those dying from ovarian-, prostate-, brain-or hematologic cancers were typically less socioeconomically deprived, with a mean SIMD category value of 3 or more., indicating less deprived areas. There was also variation in rural-urban status of patients by cancer type.

**Table 2:** Study population by cancer type

|  | All | Cancer | Bron./Lung | Col/Rect | Esoph/Sto. | Liver/Intr. | Pancr eas | Kidney/Bl. | Breast | Ovary | Prostate | Brain | Hematolo. |
| --- | --- | --- | --- | --- | --- | --- | --- | --- | --- | --- | --- | --- | --- |
|  | Mean (sd) | Mean (sd) | Mean (sd) | Mean (sd) | Mean (sd) | Mean (sd) | Mean (sd) | Mean (sd) | Mean (sd) | Mean (sd) | Mean (sd) | Mean (sd) | Mean (sd) |
| age death | 80.01 (8.15) | 78.85 (7.82) | 77.32 (7.35) | 80.15 (7.86) | 78.56 (7.76) | 78.20 (7.48) | 77.99 (7.67) | 80.01 (7.67) | 81.16 (8.57) | 78.61 (7.78) | 80.87 (7.84) | 75.61 (6.89) | 79.66 (7.57) |
| sex | 1.48 (0.50) | 1.48 (0.49) | 1.49 (0.50) | 1.48 (0.50) | 1.39 (0.48) | 1.41 (0.49) | 1.53 (0.50) | 1.40 (0.49) | 1.99 (0.10) | 2.00 (0.00) | 1.00 (0.00) | 1.42 (0.49) | 1.42 (0.49) |
| CI | 2.42 (2.17) | 2.70 (2.24) | 2.66 (2.16) | 2.76 (2.28) | 2.39 (1.55) | 2.61 (1.92) | 2.39 (1.79) | 2.66 (2.22) | 3.39 (2.93) | 3.56 (2.87) | 3.01 (2.71) | 2.16 (1.30) | 2.02 (1.34) |
| SIMD | 2.81 (1.38) | 2.79 (1.38) | 2.53 (1.33) | 2.93 (1.39) | 2.83 (1.38) | 2.81 (1.41) | 2.95 (1.38) | 2.84 (1.37) | 2.96 (1.38) | 3.00 (1.40) | 3.04 (1.38) | 3.11 (1.36) | 3.05 (1.38) |
| RU | 2.02 (1.36) | 2.02 (1.36) | 1.94 (1.27) | 2.03 (1.39) | 2.06 (1.39) | 1.95 (1.31) | 2.08 (1.43) | 2.04 (1.36) | 2.10 (1.42) | 2.03 (1.38) | 2.10 (1.48) | 2.12 (1.48) | 2.06 (1.41) |

### Inpatient, outpatient and day case use

Of the 85,732 patients included in the final analysis, 78,919 (92.05%) patients’ had at least one inpatient stay or day case activity during their last year of life, whilst 75,863 (88.49%) had at least one outpatient attendance. The number of patients with no in-or outpatient appointment was less than 0.1%. The average number of inpatient stays, length of stay per inpatient stay, number of outpatient and day case appointments are presented in Supplementary Table 1. Results are split into cancer types and presented with regards to proximity to death. Over the last year of life, patients with haematological cancers had the most inpatient appointments, with an average of 11.8 stays; but with a comparably shorter mean length of stay (LOS) of 6.1 days per stay. The longest average LOS was recorded for brain cancer patients followed by prostate cancer patients, with 9.07 and 7.94 days per stay, respectively. Haematological cancer patients had the highest number of outpatient appointments in their last year of life (mean 9.9 appointments), followed by ovary and breast cancer patients, with 6.6 and 6.3 appointments respectively. Relatively low resource use was captured for day cases, with hematological and ovarian cancer patients being most frequent day case attenders.

### Patterns of healthcare use and associated costs by cancer type

Figure 1 demonstrates significant variation in patterns of healthcare use across cancer types. Inpatient hospitalisation rates increased with proximity to death for all cancer types, though at very different rates. Varying degrees of use were observed between patients with cancer as main the cause of death and those who died from other causes, with the latter utilising fewer resources in their last year of life in all three categories (inpatient, outpatient and day case use). Once again, different patterns emerged depending on cancer type. Patients with haematological cancers were consistently high users of secondary care, with associated high costs. Considering the solid tumours only, ovarian cancer patients accessed considerably more outpatient and day care over the last year of life. Patients with certain other cancer types, for example those with brain cancer, recorded a high use of inpatient care whilst other resource use remained low. Conversely, patients with other types of cancer, such as cancers of the lower gastrointestinal tract, showed a high frequency of outpatient use whilst their use of inpatient services was minimal. Overall, frequency of outpatient care remained relatively constant over the last year of life for most cancer groups, except for those who died from haematological cancers. This patient cohort showed a steep increase in outpatient use up to the last month prior to death, followed by a sizeable drop in the last month prior to death.

**Figure 1:**
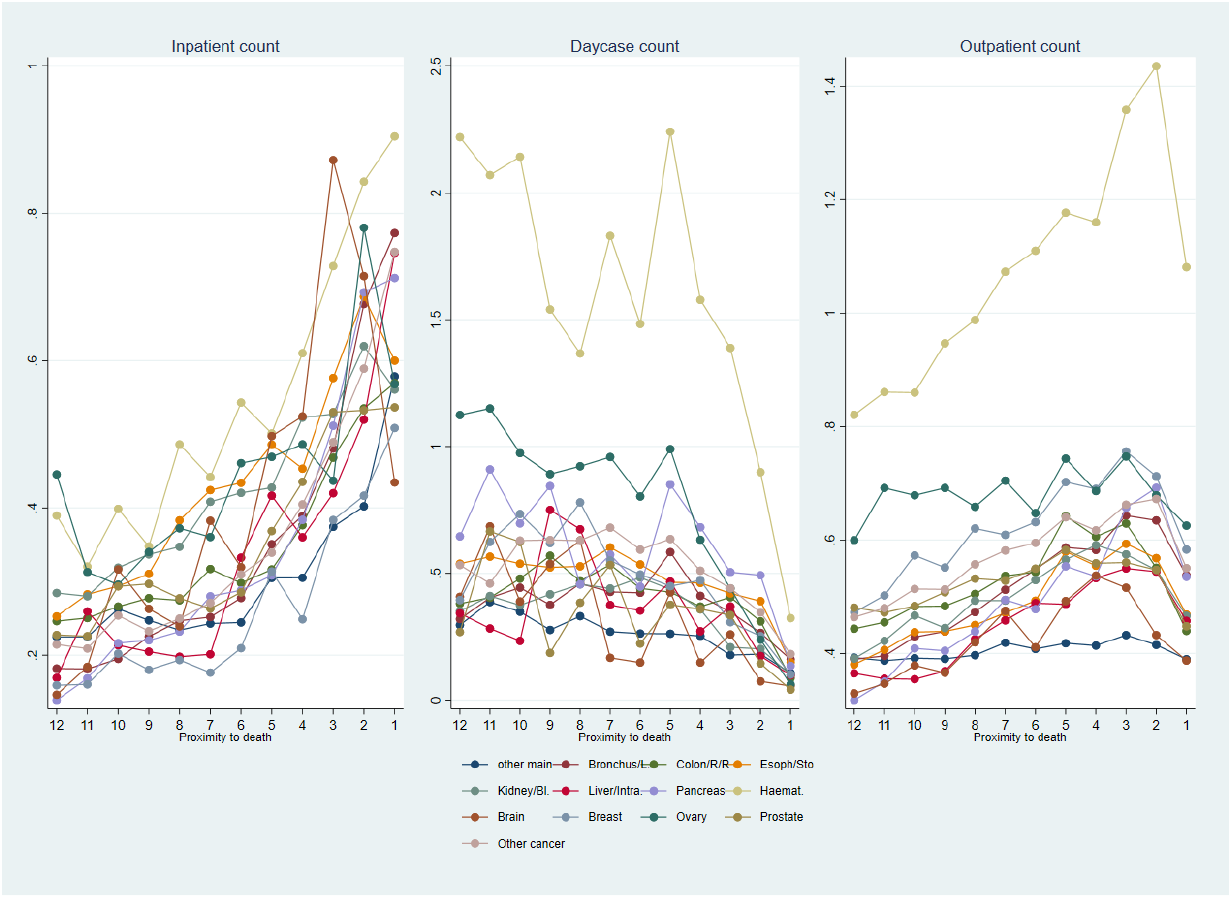
Resource use - inpatient, outpatient, day case patterns by proximity to death for individual cancer types

The results showing the frequency of resource use are in line with the corresponding costs shown in Table 4 (and Supplementary Graph 4) which confirm the outlier position of haematological cancers in terms of costs. Costs for inpatient stays in the last year of life followed a clear and consistent pattern across all cancer types, with a steep rise over the last three months of life.

### Univariate Analysis and Multivariate Analysis

Results from the univariate analysis (Supplementary Tables 2 to 6) reveal significantly lower costs associated with increased age, female gender and residing in the 3^rd^ and 4^th^ SIMD decile categories (some of the less deprived postcode areas). Costs were observed to be slightly higher for those in SIMD quintile five (least deprived category) and for those living in the most urban areas. However, the majority of the variation in costs was explained by the clinical profiles of decedents, age and their level of comorbidity.

Results of the multivariate analysis (including the sum of inpatient, outpatient and day case costs) (Table 3) confirmed the univariate results with the exception of the SIMD indicator, where only the findings related to the fifth quintile remained statistically significant. This may be due to its correlation with the rural-urban indicator. Adding in an interaction term between age and comorbidity in an attempt to unpick the effects and split the population by cause of death, it was observed that for decedents for whom cancer was the main cause of death, age and comorbidity burden had a bigger impact on costs. (Supplementary Tables 7 and 8)

**Table 3:** GLM - multivariate

|  |  |  |
| --- | --- | --- |
| 60-64 | 0 | [0,0] |
| 65-69 | -0.0845*** | [-0.108,-0.0613] |
| 70-74 | -0.159*** | [-0.182,-0.137] |
| 75-79 | -0.229*** | [-0.252,-0.207] |
| 80-84 | -0.307*** | [-0.331,-0.283] |
| 85-89 | -0.357*** | [-0.385,-0.328] |
| 90+ | -0.396*** | [-0.441,-0.352] |
| Comorbidity Index (CI) 0 or not recorded | 0 | [0,0] |
| CI 1- CI 3 | 0.661*** | [0.645,0.677] |
| CI 4 - CI 6 | 0.649*** | [0.617,0.677] |
| CI 7 - CI 12 | 0.909*** | [0.881,0.936] |
| Male | 0 | [0,0] |
| Female | -0.0589*** | [-0.0718,-0.0459] |
| SIMD 1st quintile (Most deprived) | 0 | [0,0] |
| SIMD 2<sup>nd</sup> quintile | 0.0323 | [-0.0159,0.0224] |
| SIMD 3<sup>rd</sup> quintile | -0.0193 | [-0.0394,0.000707] |
| SIMD 4<sup>th</sup> quintile | -0.0194 | [-0.0401,0.0661] |
| SIMD 5th quintile (Least deprived) | 0.0499*** | [0.0237,0.0661] |
| 1 - Large urban area | 0 | [0,0] |
| 2 - Other Urban Areas | -0.0454*** | [-0.0601,-0.0308] |
| 3 - Accessible Small Towns | -0.0736*** | [-0.0966,-0.0506] |
| 4 - Remote Small Towns | -0.0879*** | [-0.129,-0.0469] |
| 5 - Accessible Rural Areas | -0.149*** | [-0.203,-0.0755] |
| 6 - Remote Rural Areas | -0.103*** | [-0.130,-0.0755] |
| Constant | 9.069*** | [9.041,9.096] |
| Observations | 60728 |  |

95% confidence intervals in brackets
\* p < 0.05, \*\* p < 0.01, \*\*\* p < 0.001

### Variation of secondary care use between cancer types (GLM for individual cancers)

Lung cancer secondary care pathways were associated with the lowest costs in the last year of life, followed by those for people with liver cancer (see Supplementary Table 3). Costs were adjusted for age, gender and comorbidity as well as RU and SIMD. Overall, the results confirmed the findings for all cancers. It was observed that increasing age was associated with lower costs for all cancer types, albeit that the magnitude of cost reduction with increasing age varied by cancer type. When dying from oesophageal, stomach or brain cancer, the last life year was significantly less costly for women than men. An increased number of comorbidities led to a cost increase. Deprivation and rurality did not have a significant effect on costs of secondary care, with the exception of lung cancer patients where the treatment for those residing in areas that are more rural was shown to be slightly less costly. Looking at the costs in absolute terms it becomes clear that certain cancer types were significantly more expensive than others. The GLMs for the individual cancer types are available upon request though the translation of them into monetary values is presented in Table 4.

**Table 4:** GLM results as costs by cancer types

| Category | Cancer NOT main cause † | Bronchus/ Lung | Colon/Rectosig/ Rectum | Esophagus/ Stomage | Kidney/ Bladder | Liver/ Intrahepatic | Pancreas | Haematologic | Brain | Breast | Ovary | Prostate | Other cancers |
| --- | --- | --- | --- | --- | --- | --- | --- | --- | --- | --- | --- | --- | --- |
| 60-64 | 12489.7 | 12047.2 | 16924.9 | 15373.8 | 15498.8 | 13103.8 | 14551.3 | 36502.8 | 17527.5 | 14392.3 | 25440.8 | 15915 | 16773 |
| 65-69 | 12160.3** | 11537*** | 14619.7** | 14754 | 15325.6* | 12189.1 | 13206 | 29082.8 | 16538.3 | 15148.2 | 21695.9 | 13341.6 | 14426.7*** |
| 70-74 | 10901.1** | 10607.2*** | 12329.2*** | 12298.7 | 14011.7** | 10270.1** | 10829.2** | 25428.1* | 13991.7 | 11862.5** | 18225 | 12835.1 | 13660.5*** |
| 75-79 | 10782.7*** | 9975.3*** | 11063.3*** | 11253.4 | 12704.5** | 10707.5* | 9178.4** | 21420** | 11066.5 | 10185.3*** | 13266.7** | 12719.5 | 12420.5*** |
| 80-84 | 10027.5*** | 9421.6*** | 9933.5*** | 10570.2 | 11551.1** | 8782.1*** | 8640.3* | 15872.9** | 8598.1* | 8652.7*** | 11729.9* | 10919.5 | 10639*** |
| 85-89 | 8993*** | 9250.1*** | 9562.1** | 9505 | 10501.4** | 8487.3** | 7821.3* | 14211.1** | 9787.5 | 6697.4*** | 10232.9 | 9039.1 | 9666.4*** |
| 90+ | 8373.6*** | 9441.3*** | 8760.2** | 8912.8 | 9132.9** | 7073** | 7554.5 | 11717.7* | 10677.7 | 6010.5** | 8576.4 | 8186.6 | 9133*** |
| Male | 10926.3 | 10695.9 | 12680.2 | 13137.1 | 13783.7 | 10995.6 | 11282 | 24270.9 | 15424.5 | 12408.5 | NA | 12577.3 | 13599.5 |
| Female | 10171** | 10941.8 | 12133.4 | 11808.5*** | 12778.1* | 10952.6 | 11273.7 | 24425.3 | 13558.5** | 11371.2 | 18065.1 | NA | 12699.8*** |
| No Comorbidity | 9856.7 | 7242.5 | 8526 | 8068.2 | 9640.1 | 8154.5 | 7683.9 | 15875.9 | 9029.6 | 10458.3 | 9301.8 | 10459 | 12262.5 |
| CI (1 to 3) | 12846.6*** | 11209.6*** | 12779.3*** | 13017.8*** | 13811.7** | 11121.1** | 11832.1*** | 26158.1 | 15620.2* | 12176.6* | 17195.7 | 12900.6* | 14841.5*** |
| CI (4 to 6) | 17318.7*** | 12356.9* | 12242.8 | 13913.8* | 16209.1 | 12948.3* | 11170.1 | 27551.2 | 12891.7 | 13698.8 | 11815.6** | 14154.9 | 10405 |
| CI (7 to 12) | 15971.3* | 15886.2 | 17993* | 16038.6 | 17362.4 | 17133* | 15857.8 | 26107.2** | 16307.7 | 15457.5 | 24681.6 | 17407.8 | 15771.6*** |
| 1st (most deprived) | 11082.7 | 10864.9 | 12159.9 | 12906.9 | 14177.9 | 11405.7 | 11180.3 | 23633.3 | 13684.6 | 12077.5 | 18527.3 | 13212.6 | 12576.1 |
| 2nd | 10948.3 | 10885.1 | 12423.7 | 12523.1 | 13267.6* | 10874.3 | 10355.8* | 25618 | 14444.4 | 10492.5 | 18008.7 | 12304.5 | 12813.7 |
| 3rd | 10209*** | 10774.5 | 11944.4 | 12607.1 | 12963.1 | 11166.7 | 10589.8 | 22568 | 14943.2 | 11296.5 | 17754.3 | 12821.4 | 12957.6* |
| 4th | 10190.1*** | 10407.9 | 12879.5* | 12528 | 13044.5 | 10221.8 | 12213.8 | 24531.9 | 15078.7 | 10266.7 | 17673.3 | 11710* | 13205.5* |
| 5th (least deprived) | 10318.6** | 11145.8 | 12810.7 | 12644.2 | 13491.2 | 11066.3 | 12329 | 25243.4 | 14813.9 | 12929.9 | 18376.4 | 12924.9 | 14644.6*** |
| Large urban area | 10834.5 | 11302.9 | 12937.8 | 13201.3 | 13815.5 | 11734.7 | 11488.4 | 25046.4 | 15258.6 | 11367.6 | 19223.5 | 12951 | 13335.6 |
| Other Urban Areas | 10658.9 | 10477.4*** | 12384.8 | 12338** | 13053.7** | 10301** | 11137.7 | 24397.1 | 14276.1 | 11715.9 | 17468.1* | 12431.4 | 13177.7 |
| Access. Small Towns | 10552.8* | 10346.3*** | 11255.8*** | 12091.1** | 13226.2 | 10939.4 | 11188.4 | 24969.7 | 15058.8 | 12755.7 | 16053.8* | 12598.9 | 12952.9* |
| Remote Small Towns | 9562.9 | 10728.8* | 12451.2* | 11298* | 12287.6 | 8916.5 | 9634.7* | 20620.8 | 13419.2 | 7955.4** | 14700.6* | 11011.5* | 12273.4 |
| Accessible Rural Areas | 8760.5*** | 10138.2 | 10314.5* | 14156 | 12960 | 9952.1 | 10383 | 16496.9* | 13992.2 | 9834.7 | 20233.4 | 12057.9 | 11289.7* |
| Remote Rural Areas | 9367.6*** | 9998.4*** | 11274.1*** | 11797.5** | 13268.1 | 10079.8 | 11575.8 | 21420.6** | 13291.1 | 9719.8** | 16774.7 | 11793.1 | 12926.1 |
| Observations | 21071 | 18294 | 6297 | 5520 | 3542 | 2393 | 3320 | 2331 | 1154 | 2107 | 1322 | 2837 | 15050 |

\* p < 0.05, \*\* p < 0.01, \*\*\* p < 0.001

### Interaction between age and comorbidity burden

In univariable and multivariable cost analysis for age and comorbidity, increasing age was associated with lower costs whilst an increasing number of comorbidities was associated with higher costs (Table 3). In our study population, older age was negatively correlated with a higher comorbidity burden (Table 4).

Interaction terms between age and comorbidity showed considerable variation in their relationship between the cancer types, although these indicated a general tendency for comorbidities to have less impact on costs at older ages. Based on the negative coefficient on the interaction term, a higher comorbidity burden was associated with increased costs but less so with older age. Detailed results in Supplementary table 7 and in the supplementary figures 5 to 7.

## Discussion

### Main findings

There is a pervading myth that the escalating healthcare costs observed in developed nations worldwide are solely attributable to the ageing population. Instead of age, comorbidities and cancer diagnosis were identified as the main drivers of variation in cost at end of life for cancer patients over 65. However patients age remained an important factor, with reduced costs evident for those who are the very oldest. This study’s findings are confirming the results of our recent studies, showing that patients with cancer dying at an older age use considerably less health care resources in their last year of life than their younger counterparts. (Diernberger, et al., 2021; Luta, et al., Healthcare trajectories and costs in the last year of life: a retrospective primary care and hospital analysis., 2020)

A ‘Realistic Medicine’ approach has been proposed as a response to a perception of excessive futile intervention in elderly cancer populations. Whilst previous studies have confirmed lower rates of healthcare utilisation at the end of life for those with conditions such as dementia, our results show a high use of hospital-based care for the cancer population, albeit not for all types of cancer. However, clear interactions between age and tumour type are apparent, likely reflecting differential levels of treatment intensity in cancer types where the balance of harms and benefits may be more in favour of treatment. This might be complicated by differences in the composition or severity of comorbidities in different age groups, which may not be captured by the Charlson Comorbidity Index. Further studies with more detailed exploration of comorbidity data available in routine records are needed to unpick this complex relationship.

Variation in healthcare use between cancer types was most pronounced between those with haematological cancers compared with solid tumours. Within the group of patients who had solid tumours, those with ovarian cancer had the highest secondary care use and costs; an observation potentially explained by a practice of patients with advanced disease commonly receiving systemic treatment because of high response rates. This is contrasted with brain cancer decedents who had the lowest secondary care costs; and whose disease may be characterised by low rates of control, cure or response to intervention over the last year of life. It is important to note that this study included decedents only and therefore reflects pathways and outcomes for those who were not cured of their cancer by their treatment.

Alongside the treatment context differences between cancer types, there are many clinical differences in the symptoms and complications that patients’ experience, and the consequences of these for secondary care use. For instance, patients with advanced ovarian cancer commonly experience bowel obstruction or require ascites to be drained, necessitating inpatient admission. Individuals with haematological cancers typically require regular blood product support alongside their treatment, also necessitating in- and/or outpatient care. Therefore, the cancer type and its clinical manifestations, as well as the typical treatment approaches, will necessarily inform the need for secondary care interventions.

Whilst acknowledging the valuable role that hospital-based care offers many patients with cancer in their last year of life, it is also important to consider that some secondary care interventions may not offer benefit to individuals. Perceived ‘over-medicalisation’ towards the end of life has been shown to have a negative impact on patients’ and relatives’ satisfaction with care and to be linked to a lower quality of life. (Abedini, et al., 2019; Khan, Gomes,, & Higginson, 2014; Gomes, et al., 2012) There is a clear need to elicit patient and family expectations and preferences for care, and to aim for a meaningful shared decision-making approach (Realistic Medicine, 2022); arguably at all stages of the cancer journey, but especially as illness’ advances and the likelihood of benefit from acute medical care may be diminishing.

Alongside the need to align care with patients’ needs and preferences, it is also critical that only those treatments and interventions that offer a reasonable chance of benefit are offered, in order that our scarce health care resources are utilised efficiently. New, highly effective treatments for several cancer types have been very welcome, but we cannot ignore the additional financial burden of these on our already strained health care system. It is therefore ever more crucial to ensure that treatments are targeted to those who stand to benefit the most. (Vokinger, 2020; Diernberger, K. et al. 2021) As costs at the end of life are frequently included in health economic models of new cancer drugs for reimbursement submissions, this study provides data that will be of direct use for this purpose. Furthermore, improving the quality and appropriateness of care for patients in the last phase of life is a national and international priority. (Scottish Government, 2015)

This study confirms recent research showing that secondary care costs typically rise steeply in the last months of life. (Diernberger, et al., 2021) (Luta, et al., Healthcare trajectories and costs in the last year of life: a retrospective primary care and hospital analysis., 2020). These are important findings given that the majority of cancer deaths occur in hospital, despite expressed preferences ahead of time by the majority for end-of-life care at home. (Howell, et al., 2017) A recent trend for more community-based deaths of people with cancer has been observed, and this may reflect an increasing tendency towards advance care planning. An interesting finding in our study was the association between rurality and lower hospital costs, possibly reflecting proactive primary care for more rural populations, and alternative pathways to acute hospitalisation such as community hospital admission.

### Strengths and limitations of the study

This study captures healthcare data for the entire Scottish decedent population. By including routine datasets covering the whole population, there was low risk of sampling errors and selection bias along with the inclusion of exact incidence and prevalence rates. Furthermore, the administrative datasets covered several years, supporting the inclusion of data from up to five years prior to death (informed the calculation of CCI) as well as decedents over several years. In addition, learning from routine electronic health and administrative records carried no burden for the study participants.

Although the breadth and depth of Scottish administrative data was a strength, there were some notable gaps in our data. All data included originated in secondary care, albeit across inpatient, outpatient and daycase services. Primary care ‘in hours’ and ‘out of hours’ data was not available for this project, nor was data relating to social care and specialist palliative care. These are important parts of the care jigsaw, given that even with frequent hospitalisation, most patients spent most of their last year of life being cared for in the community. Furthermore, it did not allow for comparisons in secondary care use to be drawn between patients with differing degrees of primary care or specialist palliative care input; areas which are of great relevance and interest. A further limitation arose from the way the CCI was derived. CCI values were linked solely to inpatient datasets (92% had at least one inpatient appointment) leaving 8% without a CCI value. The results relating to the CCI therefore might have excluded the ‘better managed in the community’ or ‘relatively healthier’ patients with a comorbid condition that did not lead to admission or secondary outpatient care. Further data gaps related to specialist cancer treatments such as chemotherapy or radiotherapy.

It should be noted that our parallel study in England encompassed primary care data, but only for a small sample of the English national population; thus, neither study has managed complete data capture. (Luta, et al., Under review: Title: Intensity of care in cancer patients in the last year of life: a retrospective data linkage study, 2022) Future regional studies may be more likely to achieve in-depth, near whole healthcare system examination. A further limitation of this study is that we did not hear from people with advanced cancer or those close to them about their experiences of healthcare and the extent to which the care they accessed offered them meaningful benefit. Future studies should arguably incorporate a mixed methods approach, whereby routine data provides objective data relating to clinical pathways and costs, and qualitative research alongside illuminates the subjective, lived experience. It is only by examining value from both the health system and personal perspectives that we can expect to make recommendations about how resources can be optimally targeted.

### Conclusions

We have described patterns of secondary healthcare use and associated costs for all Scottish decedents with a cancer diagnosis who died between 2012 and 2017. Our headline finding is that inpatient hospitalisation accounted for the greatest proportion of costs across all cancer types, and particularly so over the last weeks of life. This end of life phase, when deteriorating health is inevitable, is a time when we might reasonably question the value of inpatient hospital care for many.

We recommend further research to identify enablers of a potential shift from secondary care to community care at the end of life. We do not know if the observed drop-off in in day-case and outpatient activity is replaced by community services such as GP contacts and community palliative care visits, or if it is simply replaced by inpatient hospital activity as people become too frail or sick to attend on an outpatient basis. It is not clear from this study, or indeed others, whether community services are adequately resourced to meet cancer patients’ needs as they deteriorate, or if they are potentially underused. It is likely that there are some cancer types which may be more readily supported in primary care than others. We require better insight into the value of the social care system and how community care can be a realistic alternative to hospital-based care if it is both resourced and accessible. Integrated health and social care in Scotland is a new reality and provides opportunity for whole system learning. (Scottish Government, 2015) Whether primary care can be seen as a substitute for secondary care or not is the rationale for a planned research project (ECi, 2022) that will delve into primary care and community data.

## Data Availability

Data are not available for sharing via application to the Scottish Public Benefits and Privacy Panel.

## Declarations

### Ethics

Approval was granted by the Scottish Public Benefit and Privacy panel (Ref: 1617-0100) for analysis within the Scottish National Research Data Safe Haven.

### Patient consent and consent for publication

Not required

### Competing Interests

The author(s) declared no potential conflicts of interest with respect to the research, authorship, and/or publication of this article.

### Funding

This work was supported by the Health Foundation (www.health.org.uk). The funders had no role in study design, data collection and analysis, decision to publish, or preparation of the manuscript.

### Authors’ contributions

KD and led the conception and design of the study supported by PH. KD led the data acquisition, conducted data management and analysis supported by EL, EG, JM and PH. KD led the data interpretation supported by EG, EL, GT XL, JB, EL and PH. KD, EL, GT and JB drafted and revised the article supported PH. All authors critically reviewed and edited the paper and approved the final version to be published.

## Acknowledgements

The authors acknowledge the support of the Electronic Data Research and Innovation Service (eDRIS) team (Public Health Scotland) for their involvement in obtaining approvals, provisioning and linking data and the use of the secure analytical platform within the National Safe Haven. Further the authors would like to thank all members of the scientific advisory board namely: Julia Riley, Sandra Campbell, Catherine Urch, Bee Wee, Harry Quilter-Pinner, Ivor Williams and Gianluca Fontana for their valuable input.

## Provenance and peer review

Not commissioned; externally peer reviewed.

## Supplementary material

**Supplementary figure S1:**
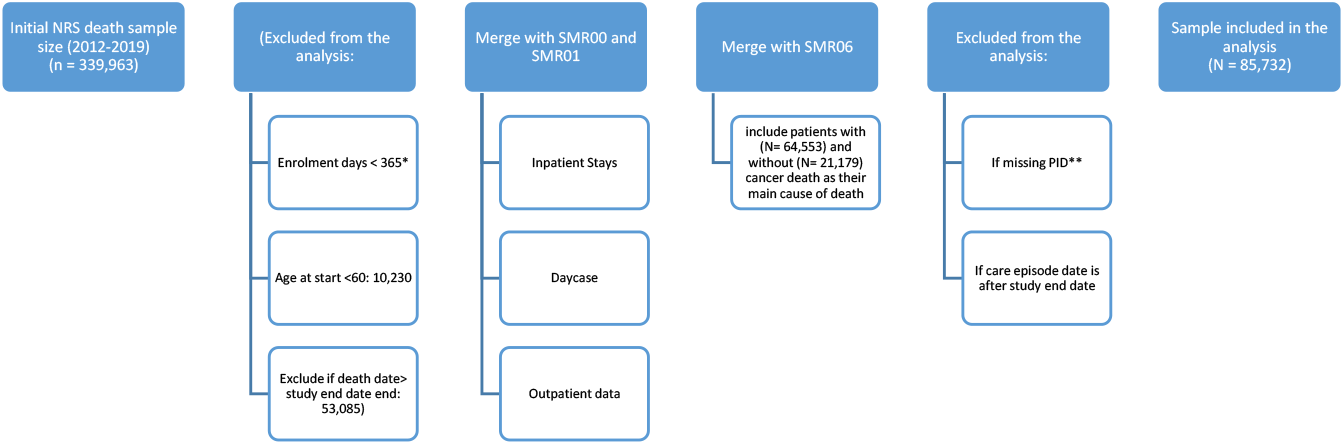
Flow chart of data linkage &inclusion/exclusion criteria *Excluded if person died within 1 year from study start date of 1^st^ Jan 2012 **PID = Person Identification Number

**Supplementary figure S2:**
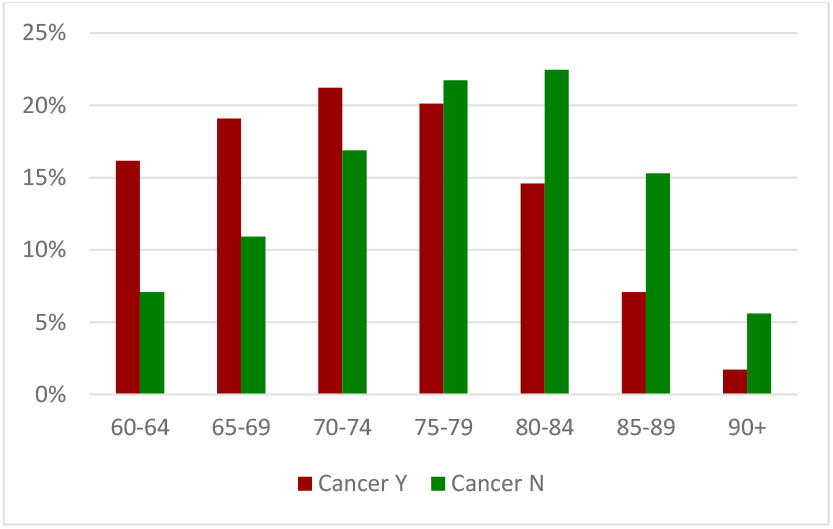
Percentage of cancer patients with cancer as their main cause of death (Cancer = Y) and “other” main death diagnosis (Cancer = N) by age

**Supplementary table 1:** Resource use (Inpatient, Inpatient LOS, Outpatient appointments and daycase use) over proximity to death (last 12, 6, 3, 1 month) by cancer type

|  | All |  | Cancer main |  | Bronchus Lung |  | Col. Rect. Rectosig. |  | Esoph. Stomache |  | Liver Intrahep. |  | Pancreas |  | Kidney Bladder |  | Breast |  | Ovary |  | Prostate |  | Brain |  | Haem. |  |
| --- | --- | --- | --- | --- | --- | --- | --- | --- | --- | --- | --- | --- | --- | --- | --- | --- | --- | --- | --- | --- | --- | --- | --- | --- | --- | --- |
|  | Mean | SD | Mean | SD | Mean | SD | Mean | SD | Mean | SD | Mean | SD | Mean | SD | Mean | SD | Mean | SD | Mean | SD | Mean | SD | Mean | SD | Mean | SD |
| Inpatient 12 | 5.88 | 5.68 | 5.99 | 5.88 | 5.30 | 4.11 | 5.75 | 4.68 | 6.15 | 4.93 | 5.21 | 4.41 | 5.68 | 5.60 | 6.36 | 4.57 | 5.60 | 5.19 | 8.41 | 8.33 | 5.72 | 4.23 | 5.27 | 3.42 | 11.80 | 14.99 |
| Average LOS 12 | 7.15 | 13.14 | 7.10 | 12.54 | 6.52 | 11.38 | 7.36 | 13.02 | 6.09 | 10.69 | 7.78 | 12.03 | 6.96 | 10.74 | 7.88 | 13.90 | 8.29 | 16.12 | 6.72 | 12.67 | 7.94 | 13.14 | 9.07 | 15.58 | 6.10 | 12.09 |
| Outpatient 12 | 5.01 | 6.00 | 5.31 | 6.28 | 5.08 | 5.04 | 5.09 | 6.35 | 4.97 | 4.64 | 4.29 | 4.65 | 4.78 | 5.86 | 4.86 | 5.00 | 6.25 | 7.34 | 6.59 | 7.08 | 5.24 | 5.14 | 3.94 | 3.87 | 9.92 | 14.31 |
| Day cases 12 | 1.15 | 4.76 | 1.36 | 5.19 | 0.97 | 3.19 | 1.19 | 3.89 | 1.45 | 4.33 | 0.64 | 2.85 | 1.52 | 5.07 | 0.92 | 3.37 | 1.40 | 4.61 | 3.20 | 8.18 | 0.83 | 2.92 | 0.30 | 1.73 | 6.45 | 15.33 |
| Inpatient 6 | 3.47 | 3.70 | 3.55 | 3.77 | 3.40 | 3.12 | 3.20 | 3.20 | 3.55 | 3.42 | 3.32 | 3.15 | 3.48 | 3.44 | 3.64 | 3.39 | 3.15 | 3.33 | 4.07 | 4.84 | 3.37 | 3.38 | 3.49 | 2.98 | 6.01 | 8.32 |
| Average LOS 6 | 7.56 | 12.31 | 7.63 | 11.98 | 7.02 | 11.07 | 8.04 | 13.02 | 6.80 | 11.16 | 8.26 | 12.14 | 7.37 | 10.40 | 8.54 | 13.13 | 8.74 | 12.84 | 7.69 | 11.35 | 8.75 | 13.56 | 9.53 | 14.42 | 6.44 | 12.10 |
| Outpatient 6 | 2.79 | 3.52 | 3.01 | 3.69 | 2.98 | 3.07 | 2.77 | 3.67 | 2.83 | 2.85 | 2.47 | 2.90 | 2.84 | 3.54 | 2.69 | 2.92 | 3.48 | 4.26 | 3.42 | 4.11 | 2.72 | 3.01 | 2.21 | 2.27 | 5.67 | 8.26 |
| Day cases 6 | 0.47 | 2.45 | 0.56 | 2.68 | 0.44 | 1.84 | 0.43 | 1.88 | 0.63 | 2.28 | 0.28 | 1.65 | 0.64 | 2.63 | 0.35 | 1.69 | 0.50 | 2.25 | 1.12 | 4.08 | 0.30 | 1.57 | 0.12 | 0.91 | 2.73 | 7.99 |
| Inpatient 3 | 2.05 | 2.50 | 2.08 | 2.50 | 2.11 | 2.31 | 1.85 | 2.28 | 2.00 | 2.45 | 2.02 | 2.32 | 2.09 | 2.31 | 2.03 | 2.45 | 1.85 | 2.24 | 2.07 | 2.67 | 1.89 | 2.39 | 1.94 | 2.36 | 2.98 | 4.40 |
| Average LOS 3 | 7.67 | 11.05 | 7.91 | 11.00 | 7.33 | 10.27 | 8.24 | 11.26 | 7.33 | 10.46 | 8.29 | 10.91 | 7.62 | 10.03 | 8.85 | 12.16 | 9.19 | 12.19 | 8.29 | 10.41 | 9.09 | 12.38 | 10.68 | 14.13 | 6.38 | 10.43 |
| Outpatient 3 | 1.19 | 1.22 | 1.28 | 1.24 | 1.34 | 1.20 | 1.13 | 1.20 | 1.25 | 1.19 | 1.12 | 1.15 | 1.27 | 1.24 | 1.17 | 1.15 | 1.38 | 1.31 | 1.40 | 1.25 | 1.12 | 1.17 | 1.01 | 1.06 | 1.82 | 1.66 |
| Day cases 3 | 0.17 | 1.13 | 0.19 | 1.24 | 0.16 | 0.91 | 0.14 | 0.90 | 0.24 | 1.16 | 0.11 | 0.92 | 0.23 | 1.24 | 0.12 | 0.81 | 0.17 | 1.09 | 0.31 | 1.64 | 0.09 | 0.67 | 0.03 | 0.37 | 0.98 | 3.72 |
| Inpatient 1 | 0.79 | 1.39 | 0.76 | 1.35 | 0.82 | 1.38 | 0.66 | 1.27 | 0.67 | 1.28 | 0.80 | 1.34 | 0.74 | 1.26 | 0.63 | 1.30 | 0.73 | 1.29 | 0.63 | 1.26 | 0.63 | 1.22 | 0.46 | 1.15 | 0.90 | 1.63 |
| Average LOS 1 | 6.01 | 6.83 | 6.45 | 7.00 | 6.14 | 6.77 | 6.60 | 7.12 | 6.33 | 6.85 | 6.66 | 7.18 | 6.66 | 7.03 | 6.88 | 6.97 | 6.64 | 7.02 | 7.33 | 7.45 | 6.98 | 7.37 | 7.98 | 8.13 | 5.26 | 6.51 |
| Outpatient 1 | 0.46 | 0.85 | 0.50 | 0.88 | 0.52 | 0.84 | 0.41 | 0.77 | 0.46 | 0.80 | 0.43 | 0.78 | 0.53 | 0.96 | 0.43 | 0.76 | 0.56 | 0.92 | 0.56 | 0.97 | 0.39 | 0.74 | 0.37 | 0.76 | 0.87 | 1.47 |
| Day cases 1 | 0.03 | 0.33 | 0.03 | 0.34 | 0.03 | 0.30 | 0.02 | 0.22 | 0.04 | 0.37 | 0.02 | 0.28 | 0.03 | 0.34 | 0.02 | 0.21 | 0.03 | 0.31 | 0.03 | 0.34 | 0.02 | 0.25 | 0.00 | 0.00 | 0.12 | 0.77 |

**Supplementary Figure 3:**
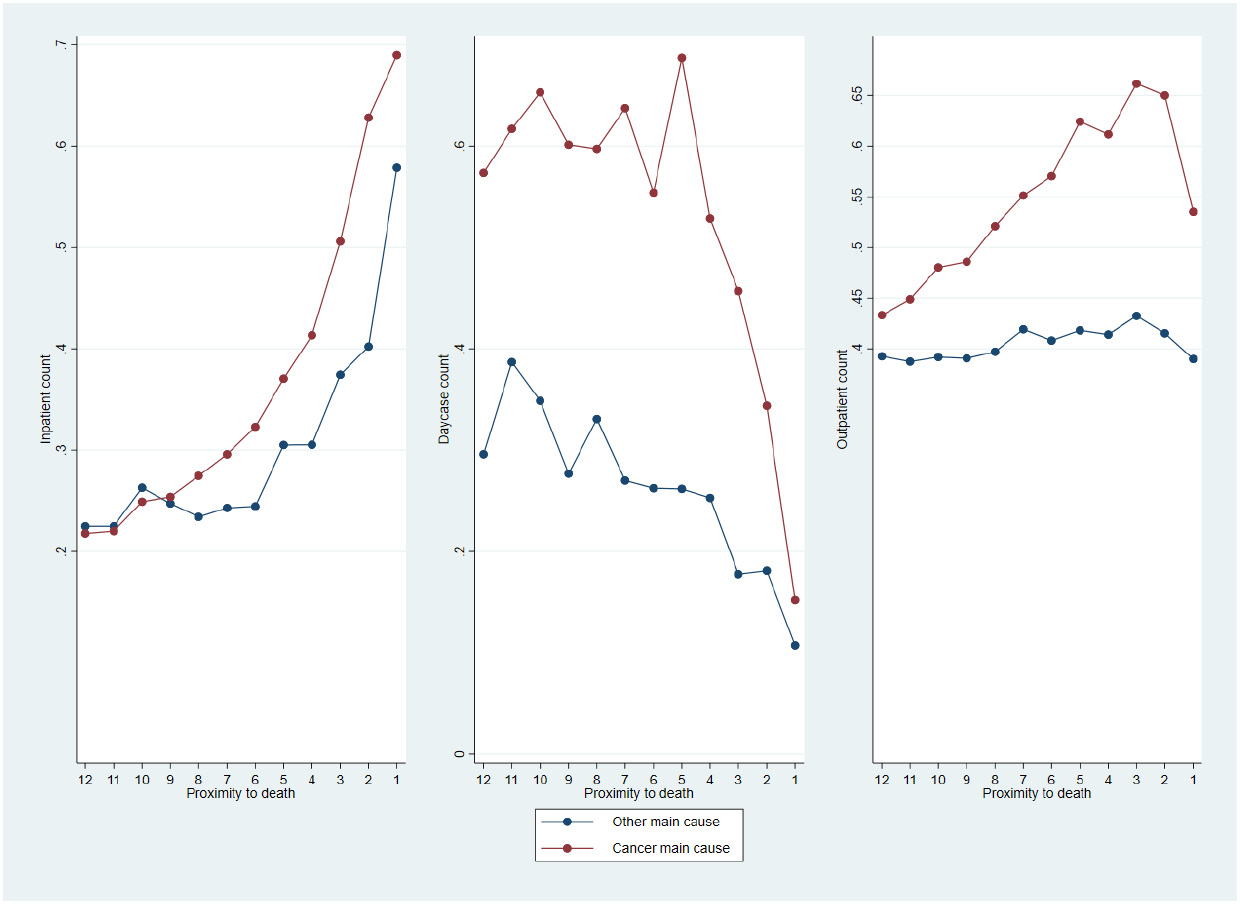
Patterns of resource use for cancer patients with and without cancer as main cause of death

**Supplementary figure 4:**
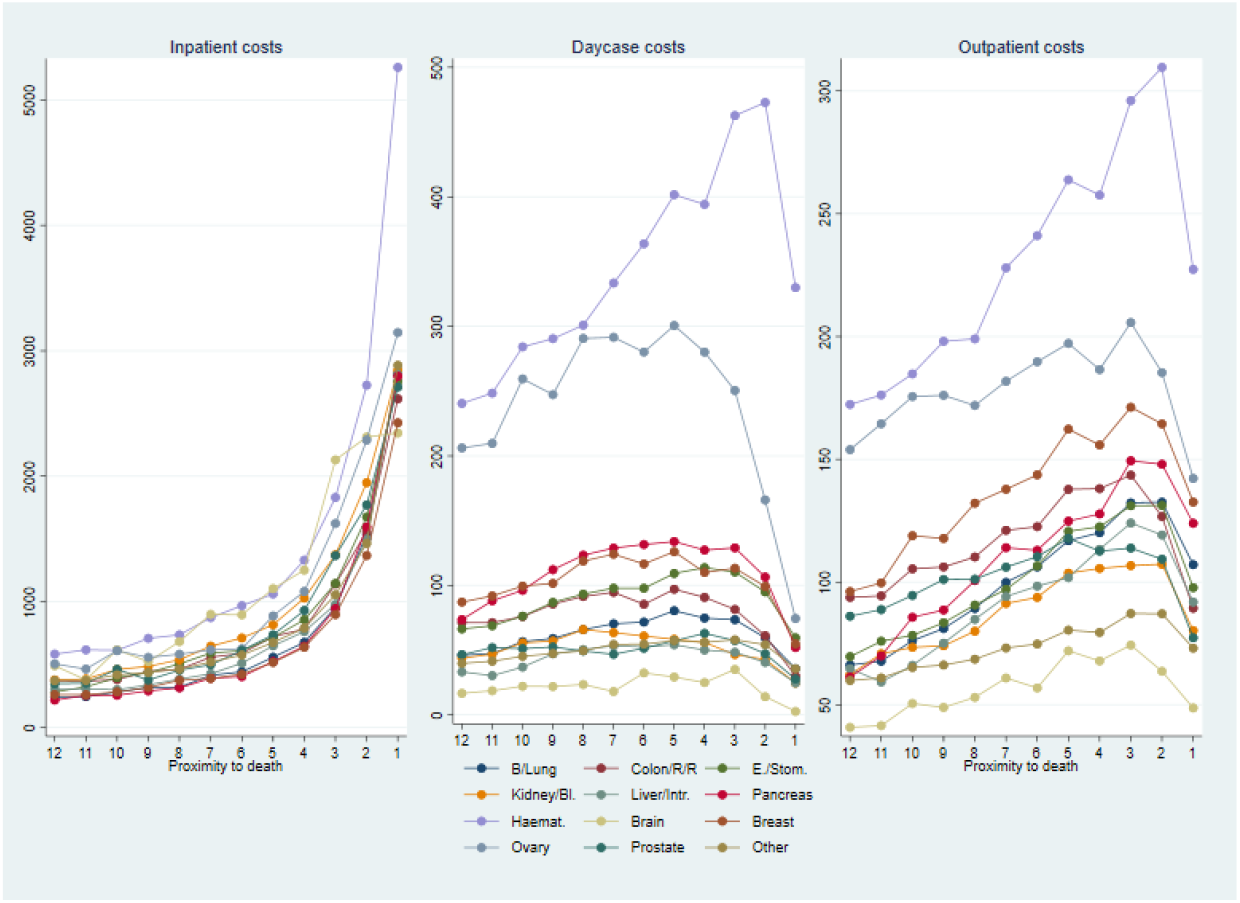
Resource use in costs - inpatient, outpatient, day case patterns by proximity to death for individual cancer type

**Supplementary table 2:** Univariate Analysis GLM - Age (margins for readability)

|  |  |  |
| --- | --- | --- |
| 60-64 | 15895.0*** | [15632.4,16157.5] |
| 65-69 | 14216.6*** | [14004.5,14428.8] |
| 70-74 | 12708.3*** | [12533.8,12882.8] |
| 75-79 | 11212.3*** | [11059.6,11365.0] |
| 80-84 | 9765.3*** | [9617.2,9913.5] |
| 85-89 | 8583.4*** | [8407.8,8759.0] |
| 90+ | 7738.8*** | [7444.4,8033.3] |
| Observations | 85328 |  |

95% confidence intervals in brackets
\* p < 0.05, \*\* p < 0.01, \*\*\* p < 0.001

**Supplementary table 3:**
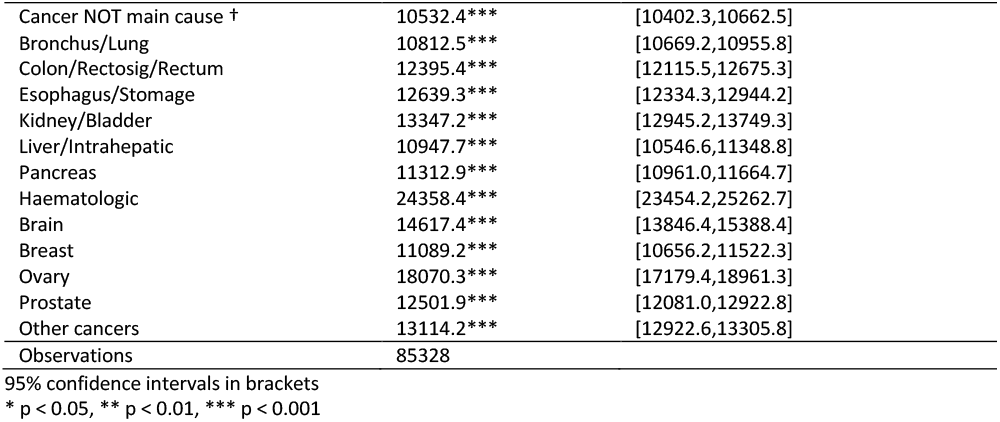
Univariate Analysis GLM - Cancertype

**Supplementary table 4:**
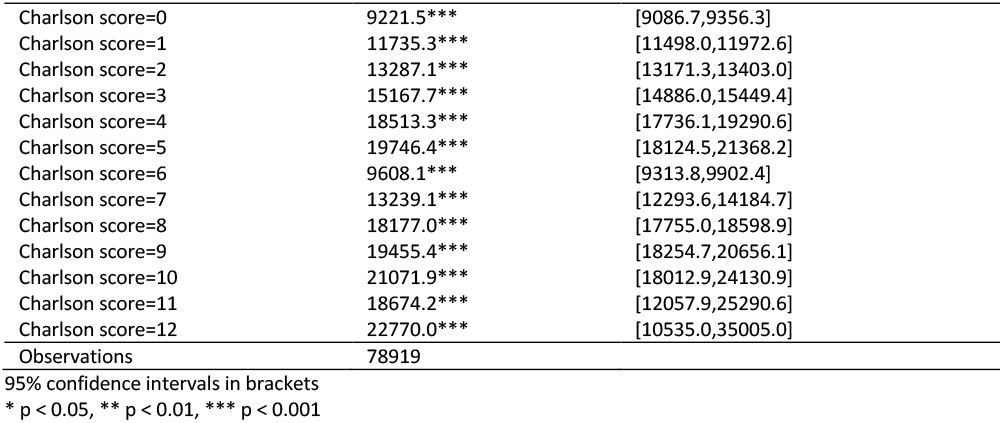
Univariate Analysis GLM - Comorbidity (Charlson score 0 to 12)

|  |  |  |
| --- | --- | --- |
| Charlson score=0 | 9221.5*** | [9086.7,9356.3] |
| Charlson score=1 | 11735.3*** | [11498.0,11972.6] |
| Charlson score=2 | 13287.1*** | [13171.3,13403.0] |
| Charlson score=3 | 15167.7*** | [14886.0,15449.4] |
| Charlson score=4 | 18513.3*** | [17736.1,19290.6] |
| Charlson score=5 | 19746.4*** | [18124.5,21368.2] |
| Charlson score=6 | 9608.1*** | [9313.8,9902.4] |
| Charlson score=7 | 13239.1*** | [12293.6,14184.7] |
| Charlson score=8 | 18177.0*** | [17755.0,18598.9] |
| Charlson score=9 | 19455.4*** | [18254.7,20656.1] |
| Charlson score=10 | 21071.9*** | [18012.9,24130.9] |
| Charlson score=11 | 18674.2*** | [12057.9,25290.6] |
| Charlson score=12 | 22770.0*** | [10535.0,35005.0] |
| Observations | 78919 |  |

95% confidence intervals in brackets
\* p < 0.05, \*\* p < 0.01, \*\*\* p < 0.001

**Supplementary table 5:** Univariate Analysis GLM - Rural-Urban Indicator

|  |  |  |
| --- | --- | --- |
| Large urban area | 12605.0*** | [12484.1,12725.9] |
| Other Urban Areas | 11867.5*** | [11740.0,11995.0] |
| Accessible Small Towns | 11717.2*** | [11475.1,11959.2] |
| Remote Small Towns | 11194.5*** | [10753.3,11635.7] |
| Accessible Rural Areas | 11005.1*** | [10431.8,11578.4] |
| Remote Rural Areas | 11296.5*** | [11017.6,11575.4] |
| Observations | 85328 |  |

95% confidence intervals in brackets
\* p < 0.05, \*\* p < 0.01, \*\*\* p < 0.001

**Supplementary table 6:** Univariate Analysis GLM - SIMD

|  |  |  |
| --- | --- | --- |
| 1st (most deprived) | 12252.7*** | [12088.3,12417.1] |
| 2nd | 12052.0*** | [11891.3,12212.7] |
| 3rd | 11815.1*** | [11649.9,11980.4] |
| 4th | 11876.8*** | [11697.1,12056.5] |
| 5th | 12645.7*** | [12442.0,12849.4] |
| Observations | 85238 |  |

95% confidence intervals in brackets
\* p < 0.05, \*\* p < 0.01, \*\*\* p < 0.001

**Supplementary table 7:** Multivariate Analysis GLM - Cancer MAIN cause of death

|  |  |  |
| --- | --- | --- |
| 60-64 | 0 | [0,0] |
| 65-69 | -0.0991*** | [-0.126,-0.0717] |
| 70-74 | -0.203*** | [-0.241,-0.165] |
| 75-79 | -0.301*** | [-0.352,-0.250] |
| 80-84 | -0.419*** | [-0.484,-0.353] |
| 85-89 | -0.484*** | [-0.565,-0.402] |
| 90+ | -0.529*** | [-0.635,-0.422] |
| CI=0 | 0 | [0,0] |
| CI=1 | 0.302*** | [0.266,0.339] |
| CI=2 | 0.187*** | [0.109,0.266] |
| CI=3 | 0.488*** | [0.371,0.604] |
| age at death | 0.00434* | [0.00100,0.00768] |
| charlson (0-12) | 0.0752*** | [0.0382,0.112] |
| age # charlson | -0.000866*** | [-0.00130,-0.000435] |
| Male | 0 | [0,0] |
| Female | -0.0366*** | [-0.0502,-0.0230] |
| 1st | 0 | [0,0] |
| 2nd | 0.00811 | [-0.0119,0.0281] |
| 3rd | 0.0216* | [0.000498,0.0426] |
| 4th | 0.0358** | [0.0141,0.0575] |
| 5th | 0.0983*** | [0.0760,0.121] |
| RU=1 | 0 | [0,0] |
| RU=2 | -0.0521*** | [-0.0675,-0.0367] |
| RU=3 | -0.0777*** | [-0.102,-0.0535] |
| RU=4 | -0.0914*** | [-0.135,-0.0480] |
| RU=5 | -0.0996*** | [-0.156,-0.0437] |
| RU=6 | -0.0789*** | [-0.108,-0.0501] |
| Constant | 9.080*** | [8.844,9.316] |
| Observations | 60728 |  |

95% confidence intervals in brackets
\* p < 0.05, \*\* p < 0.01, \*\*\* p < 0.001

**Supplementary table 8:**
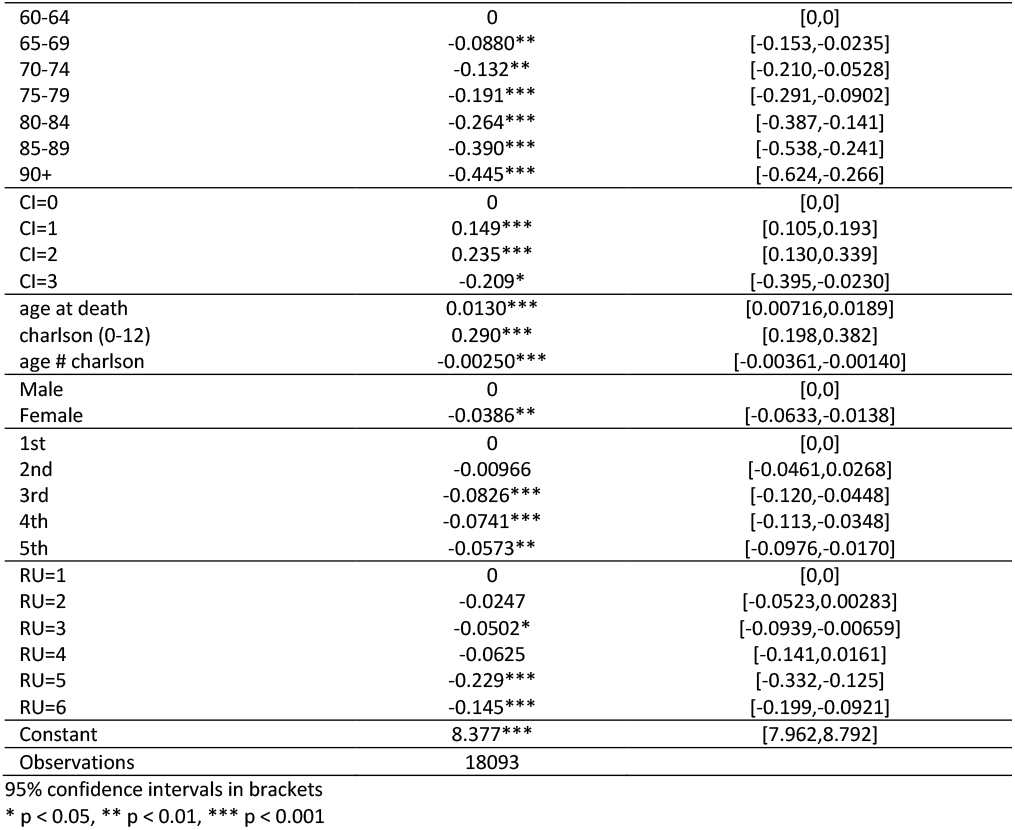
Multivariate Analysis GLM - Cancer NOT main cause of death

|  |  |  |
| --- | --- | --- |
| 60-64 | 0 | [0,0] |
| 65-69 | -0.0880** | [-0.153,-0.0235] |
| 70-74 | -0.132** | [-0.210,-0.0528] |
| 75-79 | -0.191*** | [-0.291,-0.0902] |
| 80-84 | -0.264*** | [-0.387,-0.141] |
| 85-89 | -0.390*** | [-0.538,-0.241] |
| 90+ | -0.445*** | [-0.624,-0.266] |
| CI=0 | 0 | [0,0] |
| CI=1 | 0.149*** | [0.105,0.193] |
| CI=2 | 0.235*** | [0.130,0.339] |
| CI=3 | -0.209* | [-0.395,-0.0230] |
| age at death | 0.0130*** | [0.00716,0.0189] |
| charlson (0-12) | 0.290*** | [0.198,0.382] |
| age # charlson | -0.00250*** | [-0.00361,-0.00140] |
| Male | 0 | [0,0] |
| Female | -0.0386** | [-0.0633,-0.0138] |
| 1st | 0 | [0,0] |
| 2nd | -0.00966 | [-0.0461,0.0268] |
| 3rd | -0.0826*** | [-0.120,-0.0448] |
| 4th | -0.0741*** | [-0.113,-0.0348] |
| 5th | -0.0573** | [-0.0976,-0.0170] |
| RU=1 | 0 | [0,0] |
| RU=2 | -0.0247 | [-0.0523,0.00283] |
| RU=3 | -0.0502* | [-0.0939,-0.00659] |
| RU=4 | -0.0625 | [-0.141,0.0161] |
| RU=5 | -0.229*** | [-0.332,-0.125] |
| RU=6 | -0.145*** | [-0.199,-0.0921] |
| Constant | 8.377*** | [7.962,8.792] |
| Observations | 18093 |  |

95% confidence intervals in brackets
\* p < 0.05, \*\* p < 0.01, \*\*\* p < 0.001

**Supplementary figure 5:**
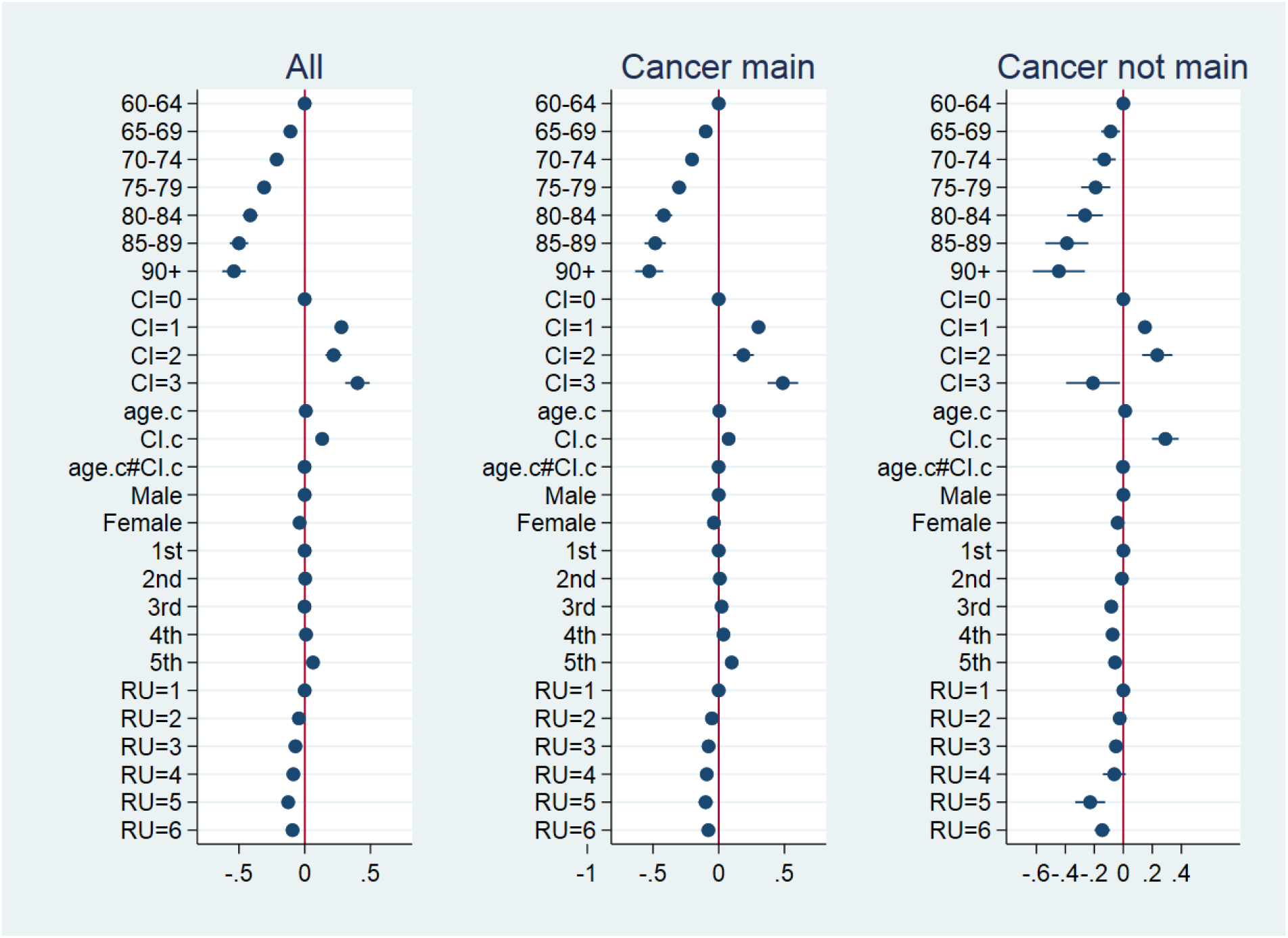
Graphical representation of GLM results including interaction term for All patients, and those with and without cancer as main cause of death

**Supplementary figure 6:**
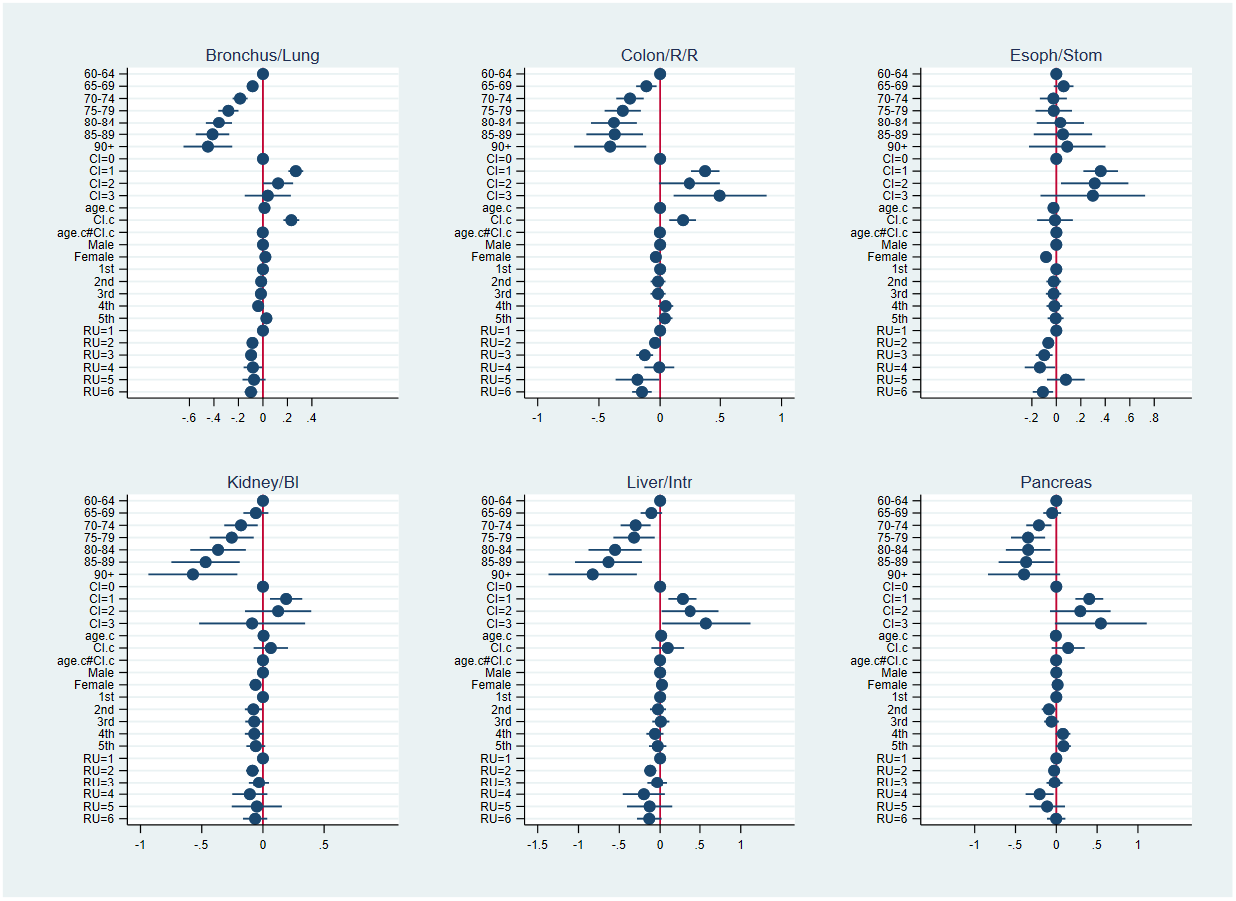
Graphical representation of GLM results including interaction term for 6 different cancer types 1/2

**Supplementary figure 7:**
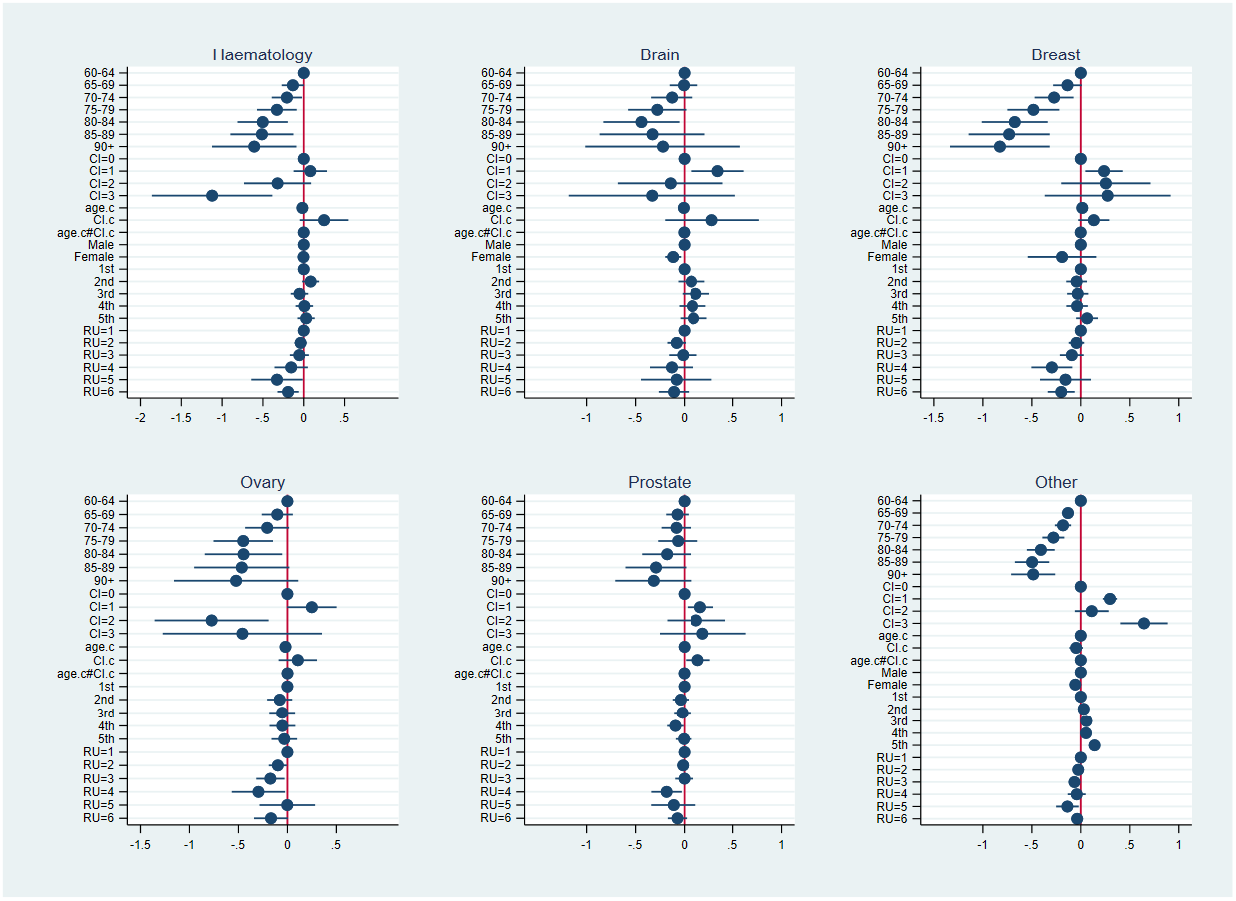
Graphical representation of GLM results including interaction term for 6 different cancer types 2/2

**Table 3:**
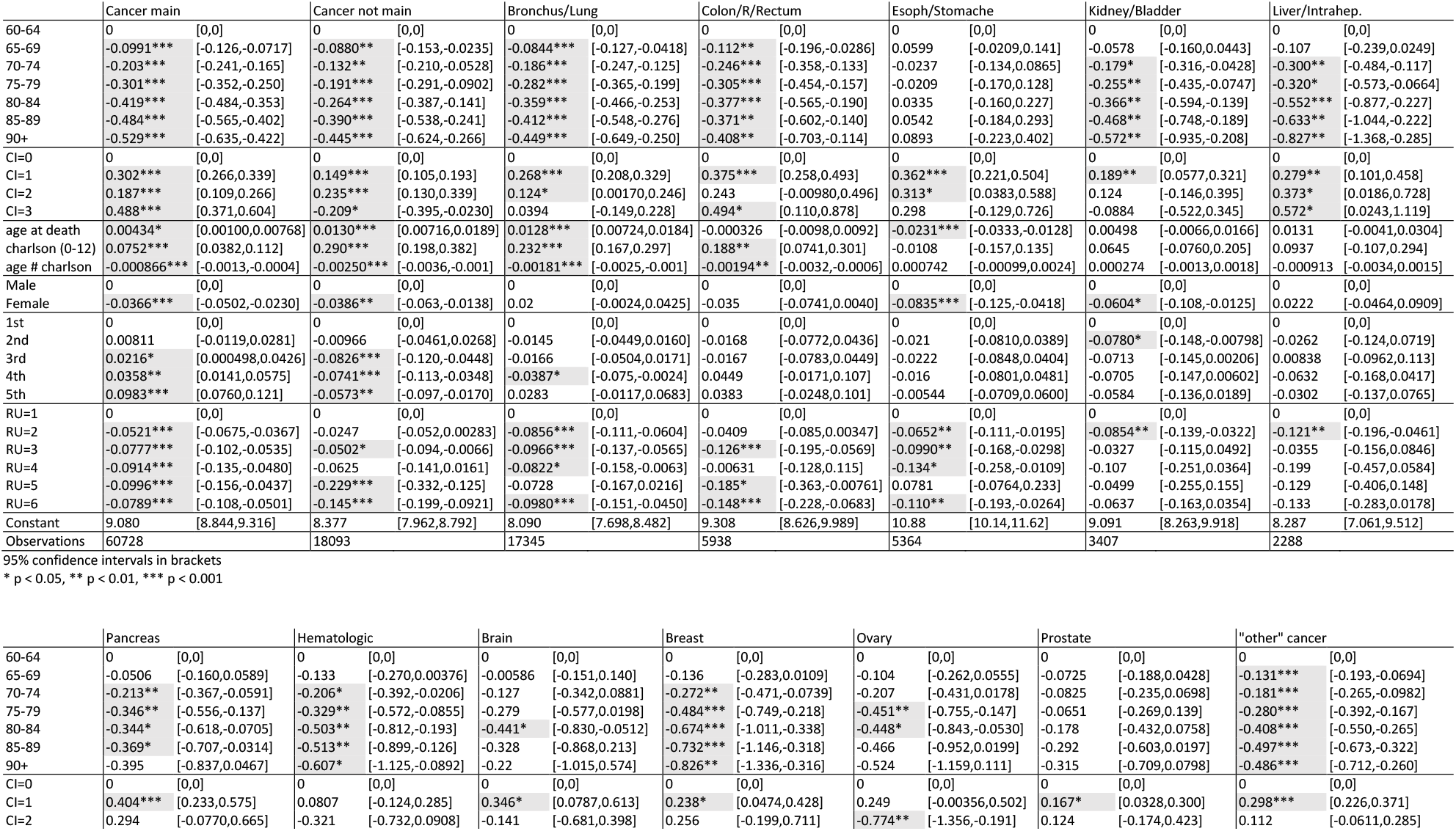
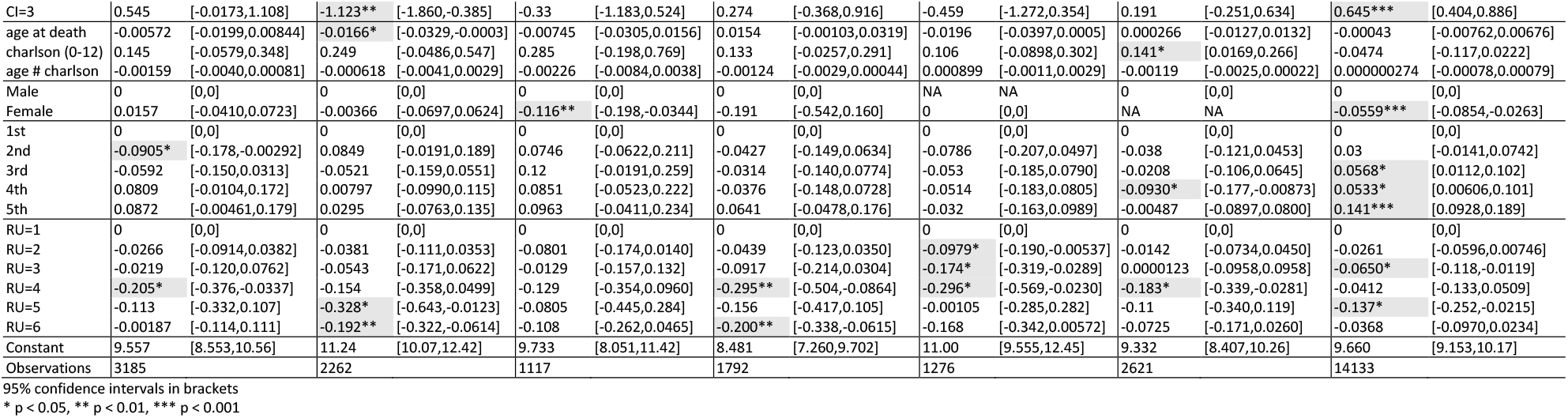
GLM results for individual cancer types

## Notes

### Competing Interest Statement

The authors have declared no competing interest.

